# Understanding Psychological Health in Adults with Irritable Bowel Syndrome: A Cross-Sectional Study Using the All of Us Research Program

**DOI:** 10.64898/2026.10.20.26365988

**Authors:** Weizi Wu, Aolan Li, Jie Chen, Eunhea You, Peiyuan Liu, Leonel Rodriguez, Ming-Hui Chen, Xiaomei Cong

## Abstract

**Background:** Psychological health is an important aspect of health and well-being among adults with irritable bowel syndrome, yet the interrelationships among individual vulnerabilities, social experiences, perceived stress, and psychological health outcomes remain poorly understood.

**Objective:** To characterize psychological health and examine direct and indirect associations of childhood adversity and adult attention-deficit/hyperactivity disorder-related symptoms within an adapted Stress Process Model.

**Design:** Cross-sectional secondary analysis.

**Setting:** The United States All of Us Research Program.

**Participants:** 2,417 adults with electronic health record-documented irritable bowel syndrome and complete data on the primary analysis variables and covariates.

**Methods:** Separate structural equation models examined childhood adversity and adult attention-deficit/hyperactivity disorder-related symptoms in relation to perceived stress, anxiety and depressive symptoms, and positive psychological well-being, with loneliness and perceived social support as psychosocial pathways. Models adjusted for sociodemographic and social-context factors.

**Results:** The median age was 54.39 years; 80.7% were female and 84.4% were White. Moderate-to-high perceived stress was reported by 56.2%, at least moderate anxiety symptoms by 21.0%, and at least moderate depressive symptoms by 24.3%. Higher ACE-8 scores were associated with greater perceived stress, anxiety symptoms, and depressive symptoms and lower psychological well-being (β = 0.063, 0.089, 0.116, and -0.100, respectively); corresponding associations for higher ASRS-6 scores were β = 0.299, 0.355, 0.390, and -0.260. Loneliness-specific indirect associations were positive for perceived stress, anxiety, and depressive symptoms (ACE-8: β = 0.023-0.034; ASRS-6: β = 0.043-0.075) and negative for psychological well-being (ACE-8: β = -0.039; ASRS-6: β = -0.089; all p < .001). Social support-specific indirect associations were not significant in the ACE-8 model and were small and significant only for anxiety and depression in the ASRS-6 model (β = 0.003-0.004).

**Conclusions:** Psychological health among adults with irritable bowel syndrome was associated with interconnected individual and social factors within an adapted Stress Process Model. Childhood adversity and adult attention-deficit/hyperactivity disorder-related symptoms showed direct and indirect associations with psychological health, with loneliness emerging as the most consistent social factor. These findings support prospective investigation of individual vulnerabilities and social experiences in psychological health. The cross-sectional design does not establish temporal ordering or causal mediation.

**What is already known:**

- Psychological distress is common in adults with irritable bowel syndrome.
- Loneliness and perceived social support describe distinct social experiences.

**What this paper adds:**

- Childhood adversity and adult attention deficit disorder with hyperactivity symptom burden were linked to psychological health.
- Loneliness was associated with distress and lower psychological well-being.

## Introduction

Irritable bowel syndrome affects approximately 6% of U.S. adults according to Rome IV criteria and is associated with substantial impairment in quality of life, daily functioning, and health care utilization ^1-3^. Although irritable bowel syndrome is defined clinically by its gastrointestinal manifestations, its recognition as a disorder of gut-brain interaction highlights the close interplay between gastrointestinal symptoms and psychological processes ^4-6^. Anxiety and depressive symptoms are common among individuals with irritable bowel syndrome, with a meta-analysis estimating prevalences of approximately 39% and 29%, respectively ^7^, and are associated with greater symptom burden, functional impairment, and poorer clinical outcomes ^8,9^. Although psychological research in irritable bowel syndrome has predominantly focused on distress, emerging evidence also highlights the relevance of positive psychological well-being to mental and physical health and quality of life ^10^. Stress is particularly relevant because it lies at the intersection of gut-brain physiology and psychological health. Stress can influence gastrointestinal function through neuroendocrine and autonomic pathways, while perceived stress reflects how individuals experience and appraise demands in their lives ^11-13^.

Understanding psychological health in adults with irritable bowel syndrome therefore requires attention to the broader individual and social contexts in which stress is experienced. The Stress Process Model provides a theoretical framework for understanding how stressors, individual characteristics, psychosocial strains, and social resources relate to psychological health ^14^. This perspective is relevant to irritable bowel syndrome because previous research has linked adverse childhood experiences and attention-deficit/hyperactivity disorder with irritable bowel syndrome ^15-18^ and has identified associations of loneliness and social relationships with irritable bowel syndrome-related outcomes ^19-21^. However, less is known about how these individual and social factors jointly relate to perceived stress and psychological health among adults with irritable bowel syndrome. Examining these factors within a shared theoretical framework may provide a more comprehensive understanding of psychological health in this population and inform future research on person-centered approaches to psychosocial assessment and symptom management.

The All of Us Research Program, a large U.S. research cohort integrating electronic health records with extensive participant-reported data ^22,23^, provides an opportunity to examine these factors together. Guided by the Stress Process Model and the measures available in All of Us, we developed an adapted conceptual framework (Supplementary Figure S1) that incorporates adverse childhood experiences as early-life adversity, adult attention-deficit/hyperactivity disorder-related symptoms as current neurobehavioral symptom burden, and loneliness and perceived social support as psychosocial experiences and resources relevant to psychological health. This study aimed to characterize psychological health among adults with irritable bowel syndrome and examine the direct and indirect associations of adverse childhood experiences and attention-deficit/hyperactivity disorder-related symptoms with perceived stress, anxiety symptoms, depressive symptoms, and positive psychological well-being through loneliness and perceived social support. Adverse childhood experiences and attention-deficit/hyperactivity disorder-related symptoms were examined in separate models. Secondary exploratory analyses incorporated personality characteristics because of their potential relevance to stress appraisal ^24,25^.

## Methods

### 2.1 Study Design and Data Source

This cross-sectional study used data from the All of Us Research Program Controlled Tier Curated Data Repository version 9 (CDRv9). The analysis integrated electronic health record data with participant-reported survey data. All analyses were conducted within the secure All of Us Researcher Workbench.

### 2.2 Study population and cohort construction

The source population included adults with available electronic health record data and sex assigned at birth recorded as female or male. Study measures were obtained from three All of Us surveys: Behavioral Health and Personality, Emotional Health History and Well-Being, and Social Determinants of Health. Because the surveys were administered at different times, eligibility was evaluated relative to each participant’s survey completion date. For each contributing survey, participants were required to be aged 18-120 years at survey completion, have completed the survey within their electronic health record observation period, and have at least 365 days of electronic health record history preceding the survey date. Irritable bowel syndrome was identified from standardized electronic health record condition records, requiring at least one qualifying record. All participants in the final analytic sample had at least one qualifying record with one of four standardized irritable bowel syndrome concepts: irritable bowel syndrome (SNOMED 10743008; OMOP 75576), irritable bowel syndrome characterized by alternating bowel habit (440544005; 4234788), irritable bowel syndrome characterized by constipation (440630006; 4261072), and irritable bowel syndrome with diarrhea (197125005; 4057826). To ensure that irritable bowel syndrome was documented before the psychosocial assessments included in the analysis, participants were required to have their first irritable bowel syndrome record on or before each of the three relevant survey completion dates. The primary analytic sample was restricted to participants with complete data on all variables included in the primary models and prespecified covariates. Figure 1 summarizes cohort construction. The survey eligibility criteria do not establish the ordering of the modeled associations. Survey timing was summarized using the earliest-to-latest survey span.

**Figure 1.**
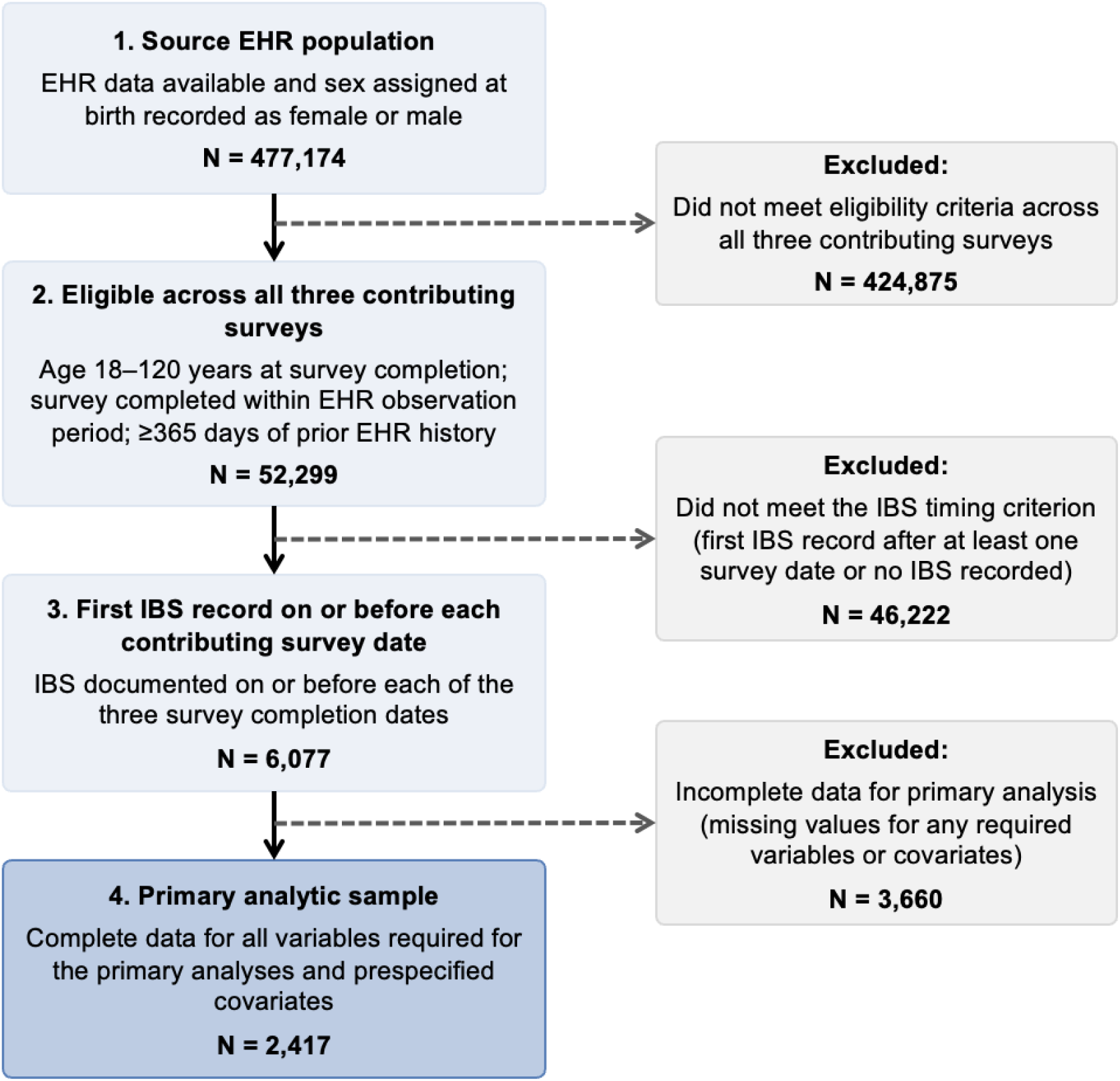
Construction of the analytic sample. Participants were required to meet age and EHR-observation criteria across all three contributing surveys and to have IBS documented on or before each survey completion date. The primary analytic sample included participants with complete data for all variables required for the primary analyses and covariates. EHR, electronic health record; IBS, irritable bowel syndrome.

### 2.3 Measures

Measures were organized according to the adapted Stress Process Model (Supplementary Figure S1), with sociodemographic and social-context factors included separately as prespecified covariates. Internal consistency of applicable multi-item scales was evaluated in the analytic sample using Cronbach’s α.

#### 2.3.1 Stress-related vulnerabilities

Two conceptually distinct constructs were examined in separate models: early-life adversity as a distal stress exposure and adult attention-deficit/hyperactivity disorder-related symptoms as an individual neurobehavioral vulnerability.

##### Early-life adversity

was assessed using eight adverse childhood experience items (ACE-8) ^26^ included in the All of Us survey. The items assessed adverse experiences occurring during childhood, including abuse and household adversity. Affirmative responses were summed to derive a cumulative score ranging from 0 to 8, with higher scores indicating exposure to more adverse childhood experiences (Cronbach’s α = .769).

##### Adult attention-deficit/hyperactivity disorder-related symptoms

were assessed using the six-item Adult ADHD Self-Report Scale (ASRS-6) ^27^. The ASRS-6 assesses the frequency of inattentive and hyperactive/impulsive symptoms in adulthood. Item responses were combined to derive a continuous symptom score ranging from 0 to 24, with higher scores indicating greater attention-deficit/hyperactivity disorder-related symptom burden. The ASRS-6 was used as a measure of symptom burden rather than to establish a clinical diagnosis of attention-deficit/hyperactivity disorder (Cronbach’s α = .820).

#### 2.3.2 Psychosocial strain and social resources

##### Loneliness

conceptualized as a psychosocial strain, was assessed using the short-form eight-item UCLA Loneliness Scale (ULS-8) ^28^ to measure feelings of loneliness and social disconnection. Item responses were summed to derive a total score ranging from 8 to 32, with higher scores indicating greater loneliness (Cronbach’s α = .883).

##### Perceived social support

conceptualized as a social resource, was assessed using the eight-item Medical Outcomes Study Social Support measure (MOS-8) ^29^. Item responses were averaged to derive a mean score ranging from 1 to 5, with higher scores indicating greater perceived social support (Cronbach’s α = .946).

#### 2.3.3 Stress appraisal and psychological health

##### Perceived stress

was assessed using the 10-item Perceived Stress Scale (PSS-10) ^30^, which evaluates the extent to which individuals perceive situations in their lives as unpredictable, uncontrollable, and overwhelming. Items were rated on a 5-point scale from 0 (never) to 4 (very often) and were summed to generate a total score ranging from 0 to 40, with higher scores indicating greater perceived stress (Cronbach’s α = .924). For descriptive purposes, scores were additionally summarized using commonly used ranges of 0-13, 14-26, and 27-40 to represent low, moderate, and high perceived stress, respectively ^31^; these categories were used descriptively and do not represent validated diagnostic thresholds.

##### Anxiety and depressive symptoms

Anxiety symptoms were assessed using the seven-item Generalized Anxiety Disorder scale (GAD-7) ^32^ with total scores ranging from 0 to 21 (Cronbach’s α = .910). Scores of 0-4, 5-9, 10-14, and 15-21 were categorized as minimal, mild, moderate, and severe anxiety symptoms, respectively. Depressive symptoms were assessed using the nine-item Patient Health Questionnaire (PHQ-9) ^33^ with total scores ranging from 0 to 27 (Cronbach’s α = .892). Scores of 0-4, 5-9, 10-14, 15-19, and 20-27 were categorized as none/minimal, mild, moderate, moderately severe, and severe depressive symptoms, respectively. Higher total scores indicated greater anxiety and depressive symptom severity, respectively.

##### Positive psychological well-being

was represented by single-item measures of happiness and perceived meaningfulness in life. Happiness responses were coded 1 = extremely happy, 2 = very happy, 3 = moderately happy, 4 = moderately unhappy, 5 = very unhappy, and 6 = extremely unhappy. Scores were reversed as 7 minus the original response so that higher scores indicated greater happiness. Meaningfulness responses were coded 1 = not at all, 2 = a little, 3 = a moderate amount, 4 = very much, and 5 = an extreme amount. Higher scores indicated greater perceived meaning in life. Happiness and meaningfulness were subsequently modeled as indicators of a latent psychological well-being factor in the structural equation models. Standardized loadings for happiness and meaningfulness were 0.806 and 0.763 in the primary ACE-8 model and 0.807 and 0.762 in the primary ASRS-6 model, respectively (Figures 2 and 3).

**Figure 2.**
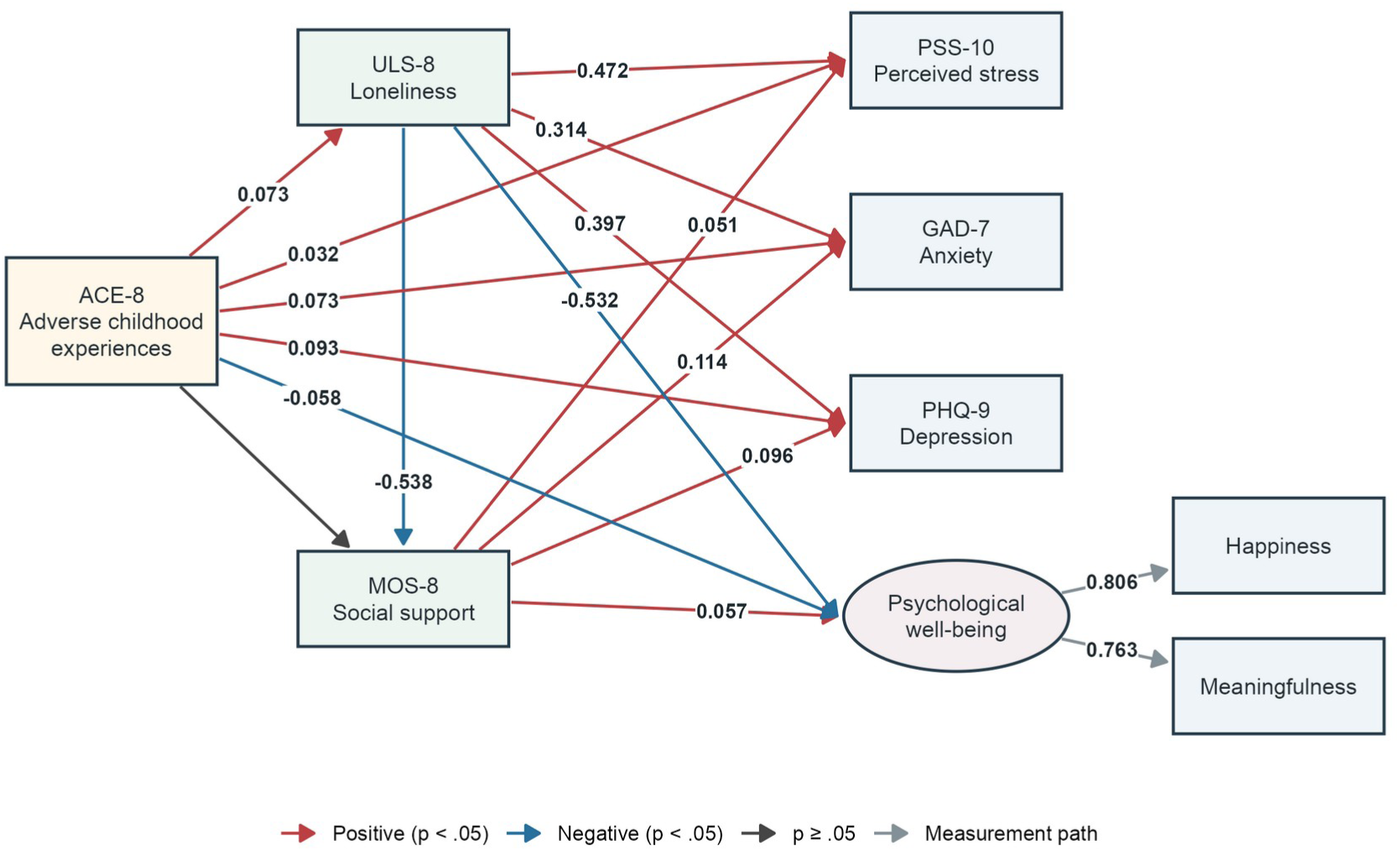
Structural equation model for early-life adversity. Fully standardized coefficients are shown for significant structural paths and for measurement loadings. Red and blue arrows indicate positive and negative associations with p < .05; dark arrows indicate p ≥ .05, and gray arrows indicate measurement loadings. Rectangles represent observed variables and the ellipse represents latent psychological well-being. N = 2,417. The model adjusted for age, sex, race/ethnicity, education, marital status, insurance, everyday discrimination, medical discrimination, and food-insecurity risk. Adjustment paths and outcome residual covariances are omitted. ACE-8, adverse childhood experiences score; ASRS-6, Adult ADHD Self-Report Scale; ULS-8, UCLA Loneliness Scale; MOS-8, Medical Outcomes Study Social Support measure; PSS-10, Perceived Stress Scale; GAD-7, Generalized Anxiety Disorder scale; PHQ-9, Patient Health Questionnaire.

**Figure 3.**
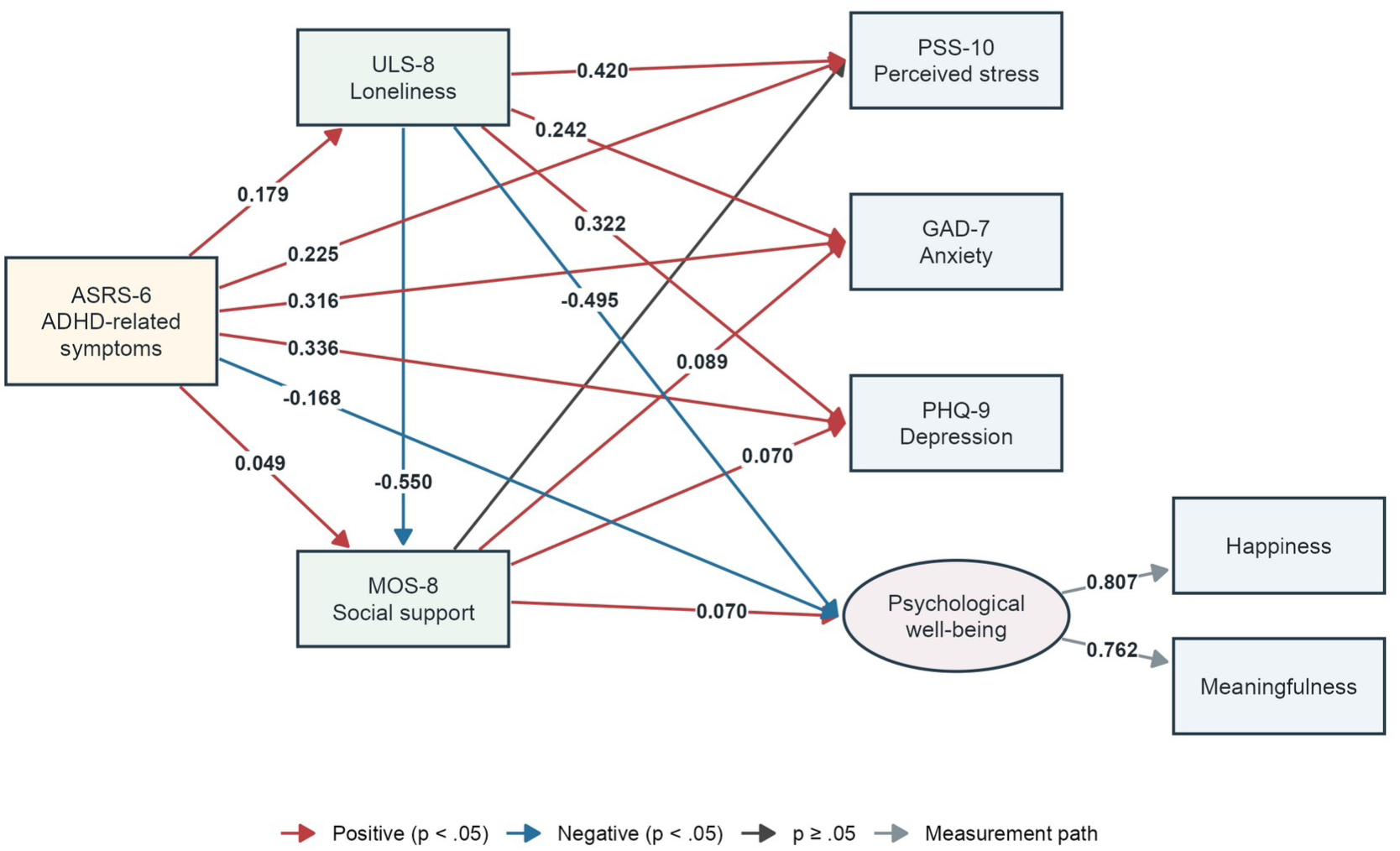
Structural equation model for adult ADHD-related symptoms. Fully standardized coefficients are shown for significant structural paths and for measurement loadings. Red and blue arrows indicate positive and negative associations with p < .05; dark arrows indicate p ≥ .05, and gray arrows indicate measurement loadings. Rectangles represent observed variables, and the ellipse represents latent psychological well-being. N = 2,417. Adjustment paths and outcome residual covariances are omitted. ACE-8, adverse childhood experiences score; ASRS-6, Adult ADHD Self-Report Scale; ULS-8, UCLA Loneliness Scale; MOS-8, Medical Outcomes Study Social Support measure; PSS-10, Perceived Stress Scale; GAD-7, Generalized Anxiety Disorder scale; PHQ-9, Patient Health Questionnaire.

#### 2.3.4 Personality characteristics

Personality characteristics were examined in exploratory secondary analyses using the 15-item Big Five Inventory-2 Extra-Short Form (BFI-2-XS) ^34^, which assesses five personality domains: extraversion, agreeableness, conscientiousness, negative emotionality, and open-mindedness. Negative emotionality was reverse-coded to derive emotional stability, such that higher scores indicated greater emotional stability. In exploratory structural equation models, the five domain scores were specified as indicators of a general personality factor to summarize their shared variation. Cronbach’s α values in the current analytic sample were .590 for extraversion, .515 for agreeableness, .671 for conscientiousness, .751 for emotional stability, and .504 for open-mindedness.

#### 2.3.5 Sociodemographic and social-context covariates

Covariates included age, sex assigned at birth, race and ethnicity, everyday discrimination, medical discrimination, food-insecurity risk, education, marital status, and health insurance. Age was determined at survey completion. Race and ethnicity were represented using the All of Us self-reported category field, derived from participants’ responses to the question, “Which categories describe you? Select all that apply.” For modeling, “More than one population” and “Other self-reported population” were combined into a “Multiracial/Other” category, whereas “Not specified” was retained separately; White was the reference category. Table 1 retains the seven original descriptive categories. Education was classified as college graduate or advanced degree versus less than a college degree; marital status as married or living with a partner versus neither; and health insurance as uninsured versus insured. Everyday discrimination was assessed using the nine-item Everyday Discrimination Scale ^35^ with scores ranging from 9 to 54, with higher scores indicating more frequent experiences of discrimination (Cronbach’s α = .910). Medical discrimination was assessed using the Everyday Discrimination Scale adapted to medical settings ^36^, with mean scores ranging from 1 to 5 and higher scores indicating greater perceived discrimination in health care settings (Cronbach’s α = .903). Food insecurity risk was assessed using the Hunger Vital Sign ^37^, and participants were classified as at risk or not at risk.

**Table 1.**
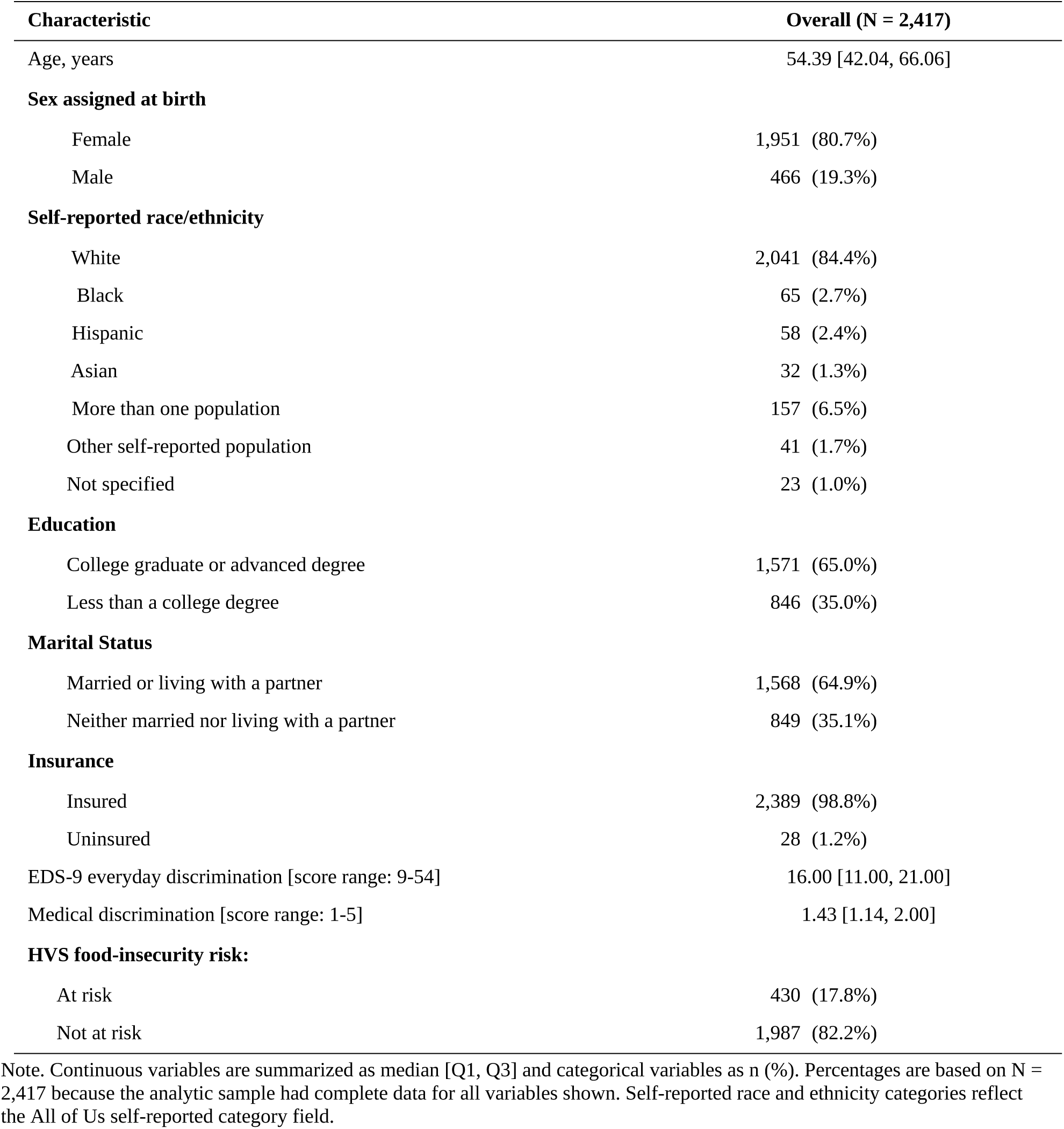
Characteristics of the analytic sample (N = 2,417)

### 2.4 Statistical analysis

Descriptive statistics used median [interquartile range (IQR)] for continuous variables and frequency (%) for categorical variables. Pearson correlations summarized pairwise associations among the 14 primary, pathway, outcome, and personality variables. Separate structural equation models were fitted for ACE-8 and ASRS-6. PSS-10 perceived stress, GAD-7 anxiety, and PHQ-9 depression were specified as three correlated observed outcomes. Positive psychological well-being was specified as a latent factor measured by happiness and meaningfulness, with equal unstandardized loadings. Each model included loneliness as a predictor of social support while retaining the conditional association between the primary exposure and social support. The primary exposure, loneliness, and social support were modeled as predictors of perceived stress, anxiety, depression, and psychological well-being. Residual covariances were freely estimated among the four outcomes. Specific indirect associations were estimated through loneliness alone, through social support alone, and through loneliness followed by social support. Specific indirect associations were calculated as products of the corresponding path coefficients, combined indirect associations as their sum, and total associations as the direct plus combined indirect association; standard errors were estimated using the delta method. Secondary models added a general personality factor measured by the five BFI-2-XS domains, an association between the primary exposure and the personality factor, and associations between the personality factor and each outcome.

All models adjusted for age, sex assigned at birth, race and ethnicity, education, marital status, health insurance, everyday discrimination, medical discrimination, and food-insecurity risk. Age was modeled using a natural spline with four degrees of freedom, and race/ethnicity were modeled categorically with White participants as the reference group. Models were estimated in lavaan using maximum likelihood with robust Huber-White standard errors and a scaled test statistic, standardized latent-variable variances, a mean structure, fixed exogenous covariates, and the specified residual covariances among outcomes ^38^. Global fit was summarized using the robust comparative fit index (CFI), Tucker-Lewis index (TLI), root mean square error of approximation (RMSEA), and standardized root mean square residual (SRMR). Fit indices were interpreted jointly rather than as pass/fail rules ^39^. Analyses were conducted using R version 4.5.3 and lavaan version 0.7-2.

Additional analyses examined the incremental variance explained by loneliness and perceived social support after accounting for each other, the exposures, and covariates. Correlation analyses and multicollinearity diagnostics were conducted to assess overlap between the two social measures (Supplementary Tables S9-S11).

### 2.5 Ethics and study registration

The All of Us Research Program was approved by the All of Us Institutional Review Board, and all participants provided informed consent for participation and use of their data for research. This secondary analysis used de-identified data in the secure Researcher Workbench and was determined to be not human subjects research by the Yale University Institutional Review Board (determination 2000043292). A project proposal was recorded in All of Us when data access was requested.

## Results

### 3.1 Sample characteristics

Of 52,299 participants meeting eligibility criteria across all three contributing surveys, 6,077 had irritable bowel syndrome documented on or before each survey date, of whom 2,417 had complete data for the primary analysis (Figure 1). Participants reported a median age of 54.39 years (IQR 42.04-66.06); 80.7% were female, 84.4% were White, and 17.8% were at risk for food insecurity. Additional sociodemographic and social-context characteristics are presented in Table 1.

The median earliest-to-latest survey span was 296 days [IQR 17-616]. Behavioral Health and Personality and Emotional Health History and Well-Being were completed on the same day by 80.8% of participants.

### 3.2 Psychological health, psychosocial characteristics, and correlations

Psychological health measures showed substantial variability across participants (Table 2). The median PSS-10 score was 15.00 (IQR 9.00-21.00); 46.2% of participants were in the moderate and 10.0% in the high perceived-stress range. Median GAD-7 and PHQ-9 scores were both 5.00 (GAD-7: IQR 2.00-8.00; PHQ-9: IQR 2.00-9.00). Overall, 21.0% of participants reported at least moderate anxiety symptoms (GAD-7 ≥10), and 24.3% reported at least moderate depressive symptoms (PHQ-9 ≥10). Median happiness and meaningfulness scores were both 4.00 (IQR 4.00-5.00 and 3.00-4.00, respectively).

**Table 2.** Descriptive statistics for the primary study measures (N = 2,417)

| Scales | Overall (N = 2,417) |
| --- | --- |
| ACE-8 [Range 0-8] | 2.00 [1.00, 4.00] |
| ASRS-6 [Range 0-24] | 9.00 [6.00, 13.00] |
| ULS-8 loneliness [Range 8-32] | 15.00 [12.00, 20.00] |
| MOS-8 social support [Range 1-5] | 4.25 [3.38, 4.88] |
| PSS-10 perceived stress [Range 0-40] | 15.00 [9.00, 21.00] |
| Low (0-13) | 1,058 (43.8%) |
| Moderate (14-26) | 1,117 (46.2%) |
| High (27-40) | 242 (10.0%) |
| GAD-7 anxiety [Range 0-21] | 5.00 [2.00, 8.00] |
| Minimal (0-4) | 1,203 (49.8%) |
| Mild (5-9) | 707 (29.3%) |
| Moderate (10-14) | 294 (12.2%) |
| Severe (15-21) | 213 (8.8%) |
| PHQ-9 depression [Range 0-27] | 5.00 [2.00, 9.00] |
| None or minimal (0-4) | 1,163 (48.1%) |
| Mild (5-9) | 666 (27.6%) |
| Moderate (10-14) | 345 (14.3%) |
| Moderately severe (15-19) | 146 (6.0%) |
| Severe (20-27) | 97 (4.0%) |
| Happiness [Range 1-6] | 4.00 [4.00, 5.00] |
| Meaningfulness [Range 1-5] | 4.00 [3.00, 4.00] |
| **Personality BFI-2-XS [Range 1-5]** |  |
| Extraversion | 3.00 [2.33, 3.67] |
| Agreeableness | 4.00 [3.67, 4.67] |
| Conscientiousness | 3.67 [3.00, 4.67] |
| Emotional Stability | 3.00 [2.00, 4.00] |
| Open-Mindedness | 4.00 [3.33, 4.67] |

**Note.** Continuous measures are presented as median [Q1, Q3], and categorical severity groups as n (%). Score ranges indicate the possible range for each measure. Emotional stability was calculated as 6 minus negative emotionality. ACE-8 = adverse childhood experiences score; ASRS-6 = Adult ADHD Self-Report Scale; BFI-2-XS = Big Five Inventory-2 Extra-Short Form; GAD-7 = Generalized Anxiety Disorder-7; MOS-8 = Medical Outcomes Study Social Support measure; PHQ-9 = Patient Health Questionnaire-9; PSS-10 = Perceived Stress Scale-10; ULS-8 = UCLA Loneliness Scale-8. PSS-10 categories are presented descriptively and do not represent validated diagnostic thresholds. GAD-7 and PHQ-9 categories reflect established symptom-severity ranges.

Participants also showed variability in stress-related and psychosocial characteristics (Table 2). Median ACE-8 and ASRS-6 scores were 2.00 (IQR 1.00-4.00) and 9.00 (IQR 6.00-13.00), respectively; median loneliness was 15.00 (IQR 12.00-20.00), and perceived social support was relatively high (median 4.25, IQR 3.38-4.88).

The correlation matrix showed consistent relationships between the psychosocial measures and psychological health (Supplementary Figure S2). Loneliness was notably correlated with perceived stress (r = 0.61), anxiety symptoms (r = 0.43), depressive symptoms (r = 0.52), happiness (r = -0.53), and meaningfulness (r = -0.50), whereas perceived social support showed associations in the opposite direction (|r| = 0.22-0.37). ACE scores showed modest correlations with psychological health measures (|r| = 0.21-0.33), while ASRS scores showed more pronounced correlations, particularly with perceived stress (r = 0.52), anxiety symptoms (r = 0.52), and depressive symptoms (r = 0.55). Loneliness and perceived social support were inversely correlated (r = -0.65; Supplementary Figure S2).

### 3.3 Early-life adversity in the adapted Stress Process Model

The primary ACE model showed good overall fit (robust CFI = 0.993, TLI = 0.954, RMSEA = 0.039 [90% CI 0.031-0.048], SRMR = 0.011) (Supplementary Table S1). Higher ACE-8 scores were associated with greater loneliness (β = 0.073, p < .001), whereas the conditional association between ACE-8 and social support was small and not statistically significant (β = -0.021, p = .212). Greater loneliness was strongly associated with lower social support (β = -0.538) and was consistently associated with poorer psychological health, including greater perceived stress (β = 0.472), anxiety symptoms (β = 0.314), and depressive symptoms (β = 0.397), and lower positive psychological well-being (β = -0.532; all p < .001). After accounting for loneliness and other model variables, higher social support showed small positive associations with perceived stress, anxiety, and depression (βs = 0.051-0.114, all p < .01), and its positive association with positive psychological well-being was statistically significant (β = 0.057, p = .044) (Figure 2; Supplementary Table S2). Across perceived stress, anxiety, and depression, adding MOS-8 after ULS-8 produced only small gains in explained variance (ΔRZ = 0.14-0.67 percentage points), whereas adding ULS-8 after MOS-8 produced larger gains (ΔRZ = 4.74-10.73 percentage points). Across both exposure models, MOS-8 coefficients for the three distress outcomes were negative when ULS-8 was omitted and positive when both measures were included. Variance inflation factors for the two social measures ranged from 1.94 to 2.15 (Supplementary Tables S9-S11).

ACE-8 remained directly associated with greater perceived stress (β = 0.032), anxiety symptoms (β = 0.073), and depressive symptoms (β = 0.093), and with lower positive psychological well-being (β = -0.058). The corresponding total associations were β = 0.063, 0.089, 0.116, and -0.100, respectively. Combined indirect associations through the psychosocial pathways were β = 0.031 for perceived stress, 0.016 for anxiety symptoms, 0.023 for depressive symptoms, and -0.042 for positive psychological well-being. Indirect associations through loneliness were statistically significant for all four outcomes, whereas indirect associations through social support alone were not. The sequential indirect associations involving loneliness followed by social support were comparatively small (Supplementary Tables S3 and S4).

### 3.4 Attention-deficit/hyperactivity disorder-related symptoms in the adapted Stress Process Model

The primary ASRS model showed good overall fit (robust CFI = 0.993, TLI = 0.954, RMSEA = 0.040 [90% CI 0.032-0.049], SRMR = 0.011). Higher ASRS-6 score was associated with greater loneliness (β = 0.179, p < .001). Greater loneliness was again associated with lower social support (β = -0.550, p < .001), while the conditional association between ASRS-6 and social support was positive and small (β = 0.049, p = .005). Across perceived stress, anxiety, and depression, adding MOS-8 after ULS-8 produced only small gains in explained variance (ΔRZ = 0.06-0.41 percentage points), whereas adding ULS-8 after MOS-8 produced larger gains (ΔRZ = 2.73-8.18 percentage points). ASRS-6 and loneliness retained statistically significant associations with perceived stress, anxiety, and depression (Figure 3; Supplementary Table S2).

Direct ASRS-6 associations were β = 0.225 for perceived stress, 0.316 for anxiety, 0.336 for depression, and -0.168 for positive psychological well-being (all p < .001). ASRS-6 total associations were β = 0.299 with perceived stress, 0.355 with anxiety, 0.390 with depression, and -0.260 with positive psychological well-being. Combined indirect associations were β = 0.074, 0.039, 0.054, and -0.092. Indirect associations through loneliness were the largest non-direct components. The positive components through social support alone and negative sequential components were small for the three psychological distress outcomes (Supplementary Tables S3 and S4).

### 3.5 Exploratory personality analyses

Exploratory models incorporating a general personality factor showed weaker fit than the primary models (ACE model: CFI = 0.910, TLI = 0.807; ASRS model: CFI = 0.886, TLI = 0.756). Across both models, emotional stability showed the strongest loading on the general personality factor (ACE model: β = 0.908; ASRS model: β = 0.833), whereas open-mindedness showed the weakest loading (β = 0.138 and 0.127, respectively) (Supplementary Figure 3; Supplementary Table S5).

In the early-life adversity model, higher ACE-8 scores were associated with lower general personality factor scores (β = -0.127, p < .001). Personality-specific indirect associations were β = 0.051 for perceived stress, 0.076 for anxiety symptoms, 0.066 for depressive symptoms, and -0.077 for psychological well-being. The corresponding total associations were β = 0.065, 0.090, 0.119, and -0.105, respectively (Supplementary Figure 3A; Supplementary Tables S6 and S7).

In the adult attention-deficit/hyperactivity disorder-related symptoms model, higher ASRS-6 scores were more strongly associated with lower scores on the general personality factor (β = -0.536, p < .001). Personality-specific indirect associations were β = 0.229 for perceived stress, 0.341 for anxiety symptoms, 0.283 for depressive symptoms, and -0.423 for psychological well-being. The corresponding total associations were β = 0.305, 0.356, 0.396, and -0.271, respectively (Supplementary Figure 3B; Supplementary Tables S6 and S7).

## Discussion

This study examined psychological health in adults with irritable bowel syndrome within a multidimensional, theory-informed framework that considered individual and social factors simultaneously. We found that 56.2% of participants reported moderate-to-high perceived stress, 21.0% reported at least moderate anxiety symptoms, and 24.3% reported at least moderate depressive symptoms. Adverse childhood experiences and attention-deficit/hyperactivity disorder-related symptoms were consistently associated with poorer psychological health across both negative and positive dimensions. Loneliness emerged as the most consistent social factor, whereas social-support-specific indirect associations were absent or comparatively small. Together, these findings suggest that understanding and addressing psychological health in adults with irritable bowel syndrome may require attention not only to individual symptoms, such as anxiety and depression, but also to broader individual and social factors, particularly experiences of social disconnection.

The associations of adverse childhood experiences with poorer psychological health are consistent with prior research linking childhood adversity with mental health across adulthood ^40^. Early adversity has also been associated with increased risk of irritable bowel syndrome ^16^, suggesting that its relevance may extend across both gastrointestinal and psychological health domains. These associations were observed in our predominantly middle-aged cohort, suggesting that retrospectively reported childhood adversity may remain relevant to psychological health later in adulthood. Although attention-deficit/hyperactivity disorder-related symptoms remain comparatively understudied in irritable bowel syndrome, their associations with psychological health are consistent with broader evidence linking adult attention-deficit/hyperactivity disorder with anxiety and depressive disorders ^41^ and attention-deficit/hyperactivity disorder-related symptoms with emotion dysregulation, a characteristic that may contribute to vulnerability to psychological distress ^42^. The presence of both direct and indirect associations further suggests that the relationships of adverse childhood experiences and attention-deficit/hyperactivity disorder-related symptoms with psychological health were not fully represented by any single psychosocial pathway examined in the models. Notably, fully standardized associations were generally larger for current attention-deficit/hyperactivity disorder-related symptom burden than for retrospectively reported childhood adversity, suggesting that neurobehavioral symptoms experienced in adulthood may be particularly relevant to concurrent psychological health in irritable bowel syndrome. This comparison should be interpreted cautiously because ACE-8 and ASRS-6 represent conceptually distinct constructs measured on different scales.

Within the two specified cross-sectional models, loneliness was associated with all four psychological health outcomes and contributed to the statistical indirect associations of both adverse childhood experiences and attention-deficit/hyperactivity disorder-related symptoms. This finding complements emerging prospective evidence linking loneliness and social disconnection with increased irritable bowel syndrome risk ^21,43^ by highlighting the relevance of loneliness to psychological health among adults with established irritable bowel syndrome. In contrast, social-support-specific indirect associations were absent in the ACE model and small in the ASRS model. Although greater perceived social support was associated with better psychological health in unadjusted analyses, several associations became small and reversed after accounting for loneliness and other factors. Previous irritable bowel syndrome research has linked greater perceived social support with lower stress, while also suggesting that negative aspects of close relationships may be more consistently associated with illness burden than supportive relationships alone ^19,20^. These adjusted associations should not be interpreted as indicating that social support is detrimental. Loneliness and perceived social support are closely related but conceptually distinct: loneliness reflects perceived inadequacy in social connection, whereas perceived social support reflects the perceived availability and adequacy of supportive resources ^44-46^. Consequently, individuals may perceive support as available while still experiencing social disconnection. When these related social experiences were considered simultaneously, subjective social disconnection provided more unique information about psychological health than perceived support availability. This distinction may be relevant to future psychosocial research and assessment, highlighting the importance of considering how socially connected individuals with irritable bowel syndrome feel, rather than only whether support is available.

The exploratory personality-expanded models should be interpreted cautiously because the five personality domains contributed unevenly to the latent general factor and overall model fit was substantially poorer than in the primary models. Emotional stability showed the strongest loading, whereas open-mindedness contributed little to the general factor, suggesting that personality may be better represented through domain-specific characteristics rather than a single general dimension in this context. This interpretation is consistent with prior irritable bowel syndrome research showing relatively robust associations of neuroticism or low emotional stability with stress, psychological distress, and symptom burden, while associations with other personality domains have been less consistent ^47-49^. Relatively stable personality characteristics may shape how individuals experience, appraise, and respond to stress, potentially contributing to differences in psychological health ^24,25^. Future studies should therefore examine individual personality domains or empirically derived profiles rather than assuming a unitary personality construct.

### Implications for nursing research and practice development

The findings highlight the relevance of individual vulnerabilities and social experiences to psychological health in adults with irritable bowel syndrome. By examining those variables within a shared theoretical framework, this study provides a basis for future research on more comprehensive, person-centered psychosocial assessment. In particular, the consistent associations of loneliness with psychological health suggest that individuals’ perceived social disconnection warrants attention alongside the availability of social support. Future longitudinal studies should examine how these factors relate to changes in psychological distress and positive well-being over time, while accounting for irritable bowel syndrome physiological symptom severity. Such evidence could inform the identification of psychosocial needs and the development of tailored supportive care approaches. Subsequent intervention studies would be needed to evaluate whether addressing these needs improves psychological and physiological health and irritable bowel syndrome-related outcomes.

### Strengths and limitations

Several strengths of this study should be considered alongside its limitations. The large All of Us cohort enabled the simultaneous examination of multiple psychosocial domains using established measures. The Stress Process Model provided a theory-informed framework for organizing these factors in relation to psychological health, although the available All of Us measures captured only selected dimensions of the framework. The primary models further accounted for key sociodemographic and social-context factors.

Nevertheless, several limitations need to be acknowledged. First, the cross-sectional design precludes establishing temporal ordering or causality; therefore, the modeled ordering should be interpreted as theory-informed, and the indirect associations should not be considered evidence of causal mediation. Second, irritable bowel syndrome was identified from electronic health records rather than through a standardized research assessment, potentially excluding individuals with undiagnosed or undocumented irritable bowel syndrome and introducing variability associated with routine clinical diagnosis. Third, the available data did not allow us to examine whether the observed psychosocial patterns varied according to current irritable bowel syndrome symptom severity. Psychosocial measures were self-reported and were not necessarily collected concurrently. Adverse childhood experiences were retrospectively reported and may be subject to recall bias, particularly given the potentially long interval since childhood in this predominantly middle-aged sample. The ASRS-6 assesses attention-deficit/hyperactivity disorder-related symptoms rather than establishing a clinical diagnosis. Fourth, the complete-case analysis excluded a substantial proportion of eligible participants and may have introduced selection bias if data completeness was associated with psychological health or other participant characteristics. The predominance of women and White participants may also limit the generalizability of the findings across more diverse populations. Finally, the poorer fit of the personality-expanded models limits the conclusions that can be drawn from these exploratory analyses. Prospective studies with repeated psychosocial and irritable bowel syndrome symptom assessments in more diverse populations are needed to clarify temporal relationships, examine associations with irritable bowel syndrome symptom burden, and assess the generalizability of these findings.

### Conclusion

Among adults with electronic health record-documented irritable bowel syndrome, adverse childhood experiences and attention-deficit/hyperactivity disorder-related symptoms were associated with perceived stress, anxiety symptoms, depressive symptoms, and psychological well-being. Loneliness contributed to indirect associations across the specified cross-sectional models, whereas social-support-specific indirect associations had confidence intervals spanning zero in the ACE-8 model and were small in the ASRS-6 model. These findings motivate prospective investigation of psychological needs in adults with irritable bowel syndrome that considers not only current psychological distress but also individual vulnerabilities and social experiences. Future longitudinal studies should clarify the temporal relationships among these factors and determine whether loneliness and related social experiences predict subsequent psychological health and irritable bowel syndrome symptom burden. Intervention studies can then evaluate whether approaches targeting social disconnection improve psychological and symptom outcomes.

## Supporting information

STROBE checklist for cross-sectional studies

## Data Availability

This study used data from the All of Us Research Program's Controlled Tier Dataset, Curated Data Repository version 9 (CDRv9; C2025Q4R6), available to authorized users through the Researcher Workbench. Access information: https://www.researchallofus.org/data-tools/workbench/.

## Acknowledgments

We gratefully acknowledge All of Us participants for their contributions, without whom this research would not have been possible. We also thank the National Institutes of Health’s All of Us Research Program for making available the participant data examined in this study.

## Funding

This work was supported in part by the National Center for Complementary and Integrative Health (NCCIH) of the National Institutes of Health under Award R3)AT012 917 (Multiple Principal Investigators: Xiaomei Cong and Jie Chen) and through a subaward from the NCCIH-funded Music Mechanisms and Technologies Network (U2)AT012 602 Subaward A25-0053-S006-A01). The content is solely the responsibility of the authors and does not necessarily represent the officcial views of the National Institutes of Health.

## Author contributions

Weizi Wu: Conceptualization, Methodology, Formal analysis, Investigation, Data curation, Visualization, Writing – original draft, Writing – review & editing.

Aolan Li: Conceptualization, Methodology, Formal analysis, Investigation, Data curation, Visualization, Writing-review & editing.

Jie Chen: Conceptualization, Validation, Funding acquisition, Writing-review & editing. Peiyuan Liu: Investigation, Data curation, Validation, Writing-review & editing.

Eunhea You: Conceptualization, Methodology, Validation, Writing-review & editing. Leonel Rodriguez: Conceptualization, Validation, Writing-review & editing.

Ming-Hui Chen: Methodology, Formal analysis, Validation, Writing-review & editing. Xiaomei Cong: Conceptualization, Supervision, Validation, Funding acquisition, Writing-review & editing.

## Declaration of competing interest

The authors declare that they have no known competing financial interests or personal relationships that could have appeared to influence the work reported in this paper.

## Declaration of generative AI and AI-assisted technologies in the manuscript preparation process

During the preparation of this work, the authors used ChatGPT (OpenAI) to assist with language refinement and clarity of the manuscript. The authors critically reviewed, verified, and edited all AI-assisted content and take full responsibility for the content of the published article.

## Notes

### Competing Interest Statement

The authors have declared no competing interest.

### Author Declarations

The Yale University Institutional Review Board determined that the present secondary analysis of de-identified data was not human subjects research (determination 2000043292; project title: NHSR: IBS AoU and UKB). The All of Us Research Program was approved by the All of Us Institutional Review Board, and all participants provided informed consent for participation and use of their data for research. Data were accessed through the secure All of Us Researcher Workbench.

