## Supplementary Tables S1-S11 and Figures S1-S3 for "Understanding Psychological Health in Adults with Irritable Bowel Syndrome: A Cross-Sectional Study Using the All of Us Research Program"

### Supplementary Table S1. Global model fit

| Model | Scaled $\chi^2$ | df | p | CFI | TLI | RMSEA (90% CI) | SRMR |
|---|---|---|---|---|---|---|---|
| ACE-8 primary model | 94.51 | 22 | <.001 | 0.993 | 0.954 | 0.039 (0.031, 0.048) | 0.011 |
| ASRS-6 primary model | 98.26 | 22 | <.001 | 0.993 | 0.954 | 0.040 (0.032, 0.049) | 0.011 |
| ACE-8 personality model | 1439.01 | 126 | <.001 | 0.910 | 0.807 | 0.067 (0.064, 0.071) | 0.038 |
| ASRS-6 personality model | 1938.75 | 126 | <.001 | 0.886 | 0.756 | 0.079 (0.076, 0.082) | 0.036 |

***Note.*** *N = 2,417 in every model. CFI, TLI, and RMSEA are robust indices. Models were estimated using MLR.*

### Supplementary Table S2. Primary structural and measurement paths

| Association | B (95% CI) | β | p |
|---|---|---|---|
| **ACE-8 primary model** | | | |
| Psychological well-being → Happiness | 0.581 (0.550, 0.612) | 0.806 | <.001 |
| Psychological well-being → Meaningfulness | 0.581 (0.550, 0.612) | 0.763 | <.001 |
| ACE-8 → Loneliness | 0.072 (0.035, 0.108) | 0.073 | <.001 |
| Loneliness → Social support | -0.514 (-0.552, -0.476) | -0.538 | <.001 |
| ACE-8 → Social support | -0.020 (-0.051, 0.011) | -0.021 | .212 |
| Loneliness → Perceived stress | 0.473 (0.432, 0.515) | 0.472 | <.001 |
| Social support → Perceived stress | 0.054 (0.014, 0.094) | 0.051 | .009 |
| ACE-8 → Perceived stress | 0.032 (0.001, 0.063) | 0.032 | .044 |
| Loneliness → Anxiety | 0.308 (0.258, 0.358) | 0.314 | <.001 |
| Social support → Anxiety | 0.117 (0.067, 0.166) | 0.114 | <.001 |
| ACE-8 → Anxiety | 0.070 (0.033, 0.108) | 0.073 | <.001 |
| Loneliness → Depression | 0.381 (0.334, 0.428) | 0.397 | <.001 |
| Social support → Depression | 0.096 (0.051, 0.142) | 0.096 | <.001 |
| ACE-8 → Depression | 0.088 (0.053, 0.123) | 0.093 | <.001 |
| Loneliness → Psychological well-being | -0.729 (-0.816, -0.643) | -0.532 | <.001 |
| Social support → Psychological well-being | 0.081 (0.002, 0.160) | 0.057 | .044 |
| ACE-8 → Psychological well-being | -0.078 (-0.136, -0.019) | -0.058 | .009 |
| **ASRS-6 primary model** | | | |
| Psychological well-being → Happiness | 0.572 (0.541, 0.603) | 0.807 | <.001 |
| Psychological well-being → Meaningfulness | 0.572 (0.541, 0.603) | 0.762 | <.001 |
| ASRS-6 → Loneliness | 0.178 (0.140, 0.217) | 0.179 | <.001 |
| Loneliness → Social support | -0.526 (-0.565, -0.487) | -0.550 | <.001 |
| ASRS-6 → Social support | 0.047 (0.015, 0.080) | 0.049 | .005 |
| Loneliness → Perceived stress | 0.421 (0.380, 0.462) | 0.420 | <.001 |
| Social support → Perceived stress | 0.036 (-0.003, 0.075) | 0.035 | .068 |
| ASRS-6 → Perceived stress | 0.225 (0.192, 0.258) | 0.225 | <.001 |
| Loneliness → Anxiety | 0.238 (0.190, 0.286) | 0.242 | <.001 |
| Social support → Anxiety | 0.092 (0.044, 0.139) | 0.089 | <.001 |
| ASRS-6 → Anxiety | 0.308 (0.270, 0.347) | 0.316 | <.001 |
| Loneliness → Depression | 0.309 (0.264, 0.354) | 0.322 | <.001 |
| Social support → Depression | 0.070 (0.027, 0.113) | 0.070 | .001 |
| ASRS-6 → Depression | 0.320 (0.284, 0.357) | 0.336 | <.001 |
| Loneliness → Psychological well-being | -0.690 (-0.778, -0.602) | -0.495 | <.001 |
| Social support → Psychological well-being | 0.102 (0.023, 0.182) | 0.070 | .012 |
| ASRS-6 → Psychological well-being | -0.233 (-0.297, -0.170) | -0.168 | <.001 |

***Note.*** *N = 2,417. B and its 95% confidence interval are in the fitted model-variable units. β is the fully standardized estimate. Intervals and two-sided p values use robust MLR inference; indirect and total associations use the delta method. Models adjusted for age using a four-degree-of-freedom natural spline, sex assigned at birth, race/ethnicity, education, marital status, insurance, everyday discrimination, medical discrimination, and food-insecurity risk.*

### Supplementary Table S3. Specific indirect associations in primary models

| Association | B (95% CI) | β | p |
|---|---|---|---|
| **ACE-8 primary model** | | | |
| Perceived stress: Via loneliness | 0.034 (0.016, 0.051) | 0.034 | <.001 |
| Perceived stress: Via social support | -0.001 (-0.003, 0.001) | -0.001 | .259 |
| Perceived stress: Via loneliness then social support | -0.002 (-0.004, 0.000) | -0.002 | .029 |
| Anxiety: Via loneliness | 0.022 (0.010, 0.034) | 0.023 | <.001 |
| Anxiety: Via social support | -0.002 (-0.006, 0.001) | -0.002 | .228 |
| Anxiety: Via loneliness then social support | -0.004 (-0.007, -0.001) | -0.004 | .004 |
| Depression: Via loneliness | 0.027 (0.013, 0.042) | 0.029 | <.001 |
| Depression: Via social support | -0.002 (-0.005, 0.001) | -0.002 | .234 |
| Depression: Via loneliness then social support | -0.004 (-0.006, -0.001) | -0.004 | .005 |
| Psychological well-being: Via loneliness | -0.052 (-0.080, -0.025) | -0.039 | <.001 |
| Psychological well-being: Via social support | -0.002 (-0.005, 0.001) | -0.001 | .289 |
| Psychological well-being: Via loneliness then social support | -0.003 (-0.006, 0.000) | -0.002 | .077 |
| **ASRS-6 primary model** | | | |
| Perceived stress: Via loneliness | 0.075 (0.058, 0.093) | 0.075 | <.001 |
| Perceived stress: Via social support | 0.002 (0.000, 0.004) | 0.002 | .119 |
| Perceived stress: Via loneliness then social support | -0.003 (-0.007, 0.000) | -0.003 | .072 |
| Anxiety: Via loneliness | 0.042 (0.030, 0.055) | 0.043 | <.001 |
| Anxiety: Via social support | 0.004 (0.001, 0.008) | 0.004 | .023 |
| Anxiety: Via loneliness then social support | -0.009 (-0.013, -0.004) | -0.009 | <.001 |
| Depression: Via loneliness | 0.055 (0.040, 0.070) | 0.058 | <.001 |
| Depression: Via social support | 0.003 (0.000, 0.006) | 0.003 | .032 |
| Depression: Via loneliness then social support | -0.007 (-0.011, -0.002) | -0.007 | .003 |
| Psychological well-being: Via loneliness | -0.123 (-0.154, -0.092) | -0.089 | <.001 |
| Psychological well-being: Via social support | 0.005 (0.000, 0.010) | 0.003 | .063 |
| Psychological well-being: Via loneliness then social support | -0.010 (-0.017, -0.002) | -0.007 | .016 |

***Note.*** *N = 2,417. B and its 95% confidence interval are in the fitted model-variable units. β is the fully standardized estimate. Intervals and two-sided p values use robust MLR inference; indirect and total associations use the delta method. Models adjusted for age using a four-degree-of-freedom natural spline, sex assigned at birth, race/ethnicity, education, marital status, insurance, everyday discrimination, medical discrimination, and food-insecurity risk.*

**Supplementary Table S4. Direct combined indirect and total associations in primary models**

| Association | B (95% CI) | β | p |
|---|---|---|---|
| **ACE-8 primary model** | | | |
| ACE-8 → Perceived stress | 0.032 (0.001, 0.063) | 0.032 | .044 |
| ACE-8 → Anxiety | 0.070 (0.033, 0.108) | 0.073 | <.001 |
| ACE-8 → Depression | 0.088 (0.053, 0.123) | 0.093 | <.001 |
| ACE-8 → Psychological well-being | -0.078 (-0.136, -0.019) | -0.058 | .009 |
| Perceived stress: Combined indirect | 0.031 (0.014, 0.047) | 0.031 | <.001 |
| Perceived stress: Total association | 0.063 (0.028, 0.097) | 0.063 | <.001 |
| Anxiety: Combined indirect | 0.015 (0.005, 0.026) | 0.016 | .003 |
| Anxiety: Total association | 0.086 (0.048, 0.124) | 0.089 | <.001 |
| Depression: Combined indirect | 0.022 (0.009, 0.035) | 0.023 | <.001 |
| Depression: Total association | 0.110 (0.073, 0.147) | 0.116 | <.001 |
| Psychological well-being: Combined indirect | -0.057 (-0.086, -0.028) | -0.042 | <.001 |
| Psychological well-being: Total association | -0.135 (-0.201, -0.069) | -0.100 | <.001 |
| **ASRS-6 primary model** | | | |
| ASRS-6 → Perceived stress | 0.225 (0.192, 0.258) | 0.225 | <.001 |
| ASRS-6 → Anxiety | 0.308 (0.270, 0.347) | 0.316 | <.001 |
| ASRS-6 → Depression | 0.320 (0.284, 0.357) | 0.336 | <.001 |
| ASRS-6 → Psychological well-being | -0.233 (-0.297, -0.170) | -0.168 | <.001 |
| Perceived stress: Combined indirect | 0.073 (0.057, 0.090) | 0.074 | <.001 |
| Perceived stress: Total association | 0.298 (0.262, 0.334) | 0.299 | <.001 |
| Anxiety: Combined indirect | 0.038 (0.027, 0.050) | 0.039 | <.001 |
| Anxiety: Total association | 0.347 (0.309, 0.385) | 0.355 | <.001 |
| Depression: Combined indirect | 0.052 (0.038, 0.065) | 0.054 | <.001 |
| Depression: Total association | 0.372 (0.336, 0.409) | 0.390 | <.001 |
| Psychological well-being: Combined indirect | -0.128 (-0.160, -0.095) | -0.092 | <.001 |
| Psychological well-being: Total association | -0.361 (-0.432, -0.291) | -0.260 | <.001 |

***Note.*** *N = 2,417. B and its 95% confidence interval are in the fitted model-variable units. β is the fully standardized estimate. Intervals and two-sided p values use robust MLR inference; indirect and total associations use the delta method. Models adjusted for age using a four-degree-of-freedom natural spline, sex assigned at birth, race/ethnicity, education, marital status, insurance, everyday discrimination, medical discrimination, and food-insecurity risk.*

### Supplementary Table S5. Measurement loadings in personality models

| Association | B (95% CI) | β | p |
|---|---|---|---|
| **ACE-8 personality model** | | | |
| Psychological well-being → Happiness | 0.454 (0.418, 0.490) | 0.787 | <.001 |
| Psychological well-being → Meaningfulness | 0.454 (0.418, 0.490) | 0.751 | <.001 |
| General personality factor → Extraversion | 0.348 (0.311, 0.384) | 0.403 | <.001 |
| General personality factor → Agreeableness | 0.221 (0.185, 0.258) | 0.259 | <.001 |
| General personality factor → Conscientiousness | 0.428 (0.394, 0.462) | 0.506 | <.001 |
| General personality factor → Emotional stability | 0.765 (0.728, 0.802) | 0.908 | <.001 |
| General personality factor → Open-mindedness | 0.115 (0.077, 0.152) | 0.138 | <.001 |
| **ASRS-6 personality model** | | | |
| Psychological well-being → Happiness | 0.415 (0.373, 0.457) | 0.790 | <.001 |
| Psychological well-being → Meaningfulness | 0.415 (0.373, 0.457) | 0.753 | <.001 |
| General personality factor → Extraversion | 0.273 (0.232, 0.314) | 0.389 | <.001 |
| General personality factor → Agreeableness | 0.175 (0.141, 0.209) | 0.253 | <.001 |
| General personality factor → Conscientiousness | 0.405 (0.381, 0.429) | 0.590 | <.001 |
| General personality factor → Emotional stability | 0.570 (0.513, 0.627) | 0.833 | <.001 |
| General personality factor → Open-mindedness | 0.085 (0.051, 0.120) | 0.127 | <.001 |

***Note.*** *N = 2,417. B and its 95% confidence interval are in the fitted model-variable units. β is the fully standardized estimate. Intervals and two-sided p values use robust MLR inference; indirect and total associations use the delta method. Models adjusted for age using a four-degree-of-freedom natural spline, sex assigned at birth, race/ethnicity, education, marital status, insurance, everyday discrimination, medical discrimination, and food-insecurity risk.*

### Supplementary Table S6. Direct combined indirect and total associations in personality models

| Association | B (95% CI) | β | p |
|---|---|---|---|
| **ACE-8 personality model** | | | |
| ACE-8 → Perceived stress | -0.008 (-0.037, 0.020) | -0.009 | .568 |
| ACE-8 → Anxiety | 0.011 (-0.022, 0.044) | 0.012 | .512 |
| ACE-8 → Depression | 0.039 (0.007, 0.070) | 0.042 | .016 |
| ACE-8 → Psychological well-being | 0.003 (-0.064, 0.070) | 0.002 | .919 |
| Perceived stress: Combined indirect | 0.071 (0.047, 0.095) | 0.074 | <.001 |
| Perceived stress: Total association | 0.063 (0.028, 0.097) | 0.065 | <.001 |
| Anxiety: Combined indirect | 0.075 (0.049, 0.101) | 0.078 | <.001 |
| Anxiety: Total association | 0.086 (0.048, 0.124) | 0.090 | <.001 |
| Depression: Combined indirect | 0.071 (0.047, 0.095) | 0.077 | <.001 |
| Depression: Total association | 0.110 (0.073, 0.147) | 0.119 | <.001 |
| Psychological well-being: Combined indirect | -0.176 (-0.235, -0.117) | -0.107 | <.001 |
| Psychological well-being: Total association | -0.173 (-0.258, -0.088) | -0.105 | <.001 |
| **ASRS-6 personality model** | | | |
| ASRS-6 → Perceived stress | 0.020 (-0.037, 0.076) | 0.020 | .496 |
| ASRS-6 → Anxiety | 0.006 (-0.059, 0.070) | 0.006 | .866 |
| ASRS-6 → Depression | 0.077 (0.016, 0.139) | 0.082 | .014 |
| ASRS-6 → Psychological well-being | 0.388 (0.204, 0.572) | 0.211 | <.001 |
| Perceived stress: Combined indirect | 0.278 (0.224, 0.332) | 0.284 | <.001 |
| Perceived stress: Total association | 0.298 (0.262, 0.334) | 0.305 | <.001 |
| Anxiety: Combined indirect | 0.341 (0.281, 0.401) | 0.351 | <.001 |
| Anxiety: Total association | 0.347 (0.309, 0.385) | 0.356 | <.001 |
| Depression: Combined indirect | 0.295 (0.239, 0.352) | 0.314 | <.001 |
| Depression: Total association | 0.372 (0.336, 0.409) | 0.396 | <.001 |
| Psychological well-being: Combined indirect | -0.886 (-1.100, -0.672) | -0.482 | <.001 |
| Psychological well-being: Total association | -0.498 (-0.602, -0.395) | -0.271 | <.001 |

***Note.*** *N = 2,417. B and its 95% confidence interval are in the fitted model-variable units. β is the fully standardized estimate. Intervals and two-sided p values use robust MLR inference; indirect and total associations use the delta method. Models adjusted for age using a four-degree-of-freedom natural spline, sex assigned at birth, race/ethnicity, education, marital status, insurance, everyday discrimination, medical discrimination, and food-insecurity risk.*

### Supplementary Table S7. Personality pathways and specific indirect associations

| Association | B (95% CI) | β | p |
|---|---|---|---|
| **ACE-8 personality model** | | | |
| ACE-8 → General personality factor | -0.150 (-0.201, -0.099) | -0.127 | <.001 |
| Perceived stress: Via loneliness | 0.022 (0.010, 0.034) | 0.023 | <.001 |
| Perceived stress: Via social support | 0.000 (-0.001, 0.001) | 0.000 | .726 |
| Perceived stress: Via loneliness then social support | 0.000 (-0.002, 0.001) | 0.000 | .716 |
| Perceived stress: Via personality | 0.049 (0.032, 0.067) | 0.051 | <.001 |
| Anxiety: Via loneliness | 0.004 (0.000, 0.009) | 0.005 | .036 |
| Anxiety: Via social support | -0.001 (-0.003, 0.001) | -0.001 | .285 |
| Anxiety: Via loneliness then social support | -0.002 (-0.004, 0.000) | -0.002 | .072 |
| Anxiety: Via personality | 0.073 (0.048, 0.098) | 0.076 | <.001 |
| Depression: Via loneliness | 0.013 (0.005, 0.020) | 0.014 | <.001 |
| Depression: Via social support | -0.001 (-0.002, 0.001) | -0.001 | .305 |
| Depression: Via loneliness then social support | -0.001 (-0.003, 0.000) | -0.002 | .095 |
| Depression: Via personality | 0.060 (0.040, 0.081) | 0.066 | <.001 |
| Psychological well-being: Via loneliness | -0.036 (-0.056, -0.016) | -0.022 | <.001 |
| Psychological well-being: Via social support | -0.004 (-0.012, 0.003) | -0.003 | .228 |
| Psychological well-being: Via loneliness then social support | -0.008 (-0.014, -0.003) | -0.005 | .004 |
| Psychological well-being: Via personality | -0.127 (-0.174, -0.081) | -0.077 | <.001 |
| **ASRS-6 personality model** | | | |
| ASRS-6 → General personality factor | -0.789 (-0.919, -0.659) | -0.536 | <.001 |
| Perceived stress: Via loneliness | 0.054 (0.040, 0.068) | 0.055 | <.001 |
| Perceived stress: Via social support | 0.000 (-0.001, 0.002) | 0.000 | .753 |
| Perceived stress: Via loneliness then social support | -0.001 (-0.004, 0.003) | -0.001 | .752 |
| Perceived stress: Via personality | 0.224 (0.173, 0.276) | 0.229 | <.001 |
| Anxiety: Via loneliness | 0.012 (0.002, 0.021) | 0.012 | .015 |
| Anxiety: Via social support | 0.002 (0.000, 0.005) | 0.002 | .104 |
| Anxiety: Via loneliness then social support | -0.004 (-0.009, 0.000) | -0.005 | .047 |
| Anxiety: Via personality | 0.332 (0.272, 0.391) | 0.341 | <.001 |
| Depression: Via loneliness | 0.030 (0.019, 0.041) | 0.032 | <.001 |
| Depression: Via social support | 0.002 (-0.001, 0.004) | 0.002 | .156 |
| Depression: Via loneliness then social support | -0.003 (-0.007, 0.001) | -0.003 | .107 |

| **Association** | **B (95% CI)** | **β** | **p** |
|---|---|---|---|
| Depression: Via personality | 0.266 (0.211, 0.322) | 0.283 | <.001 |
| Psychological well-being: Via loneliness | -0.098 (-0.127, -0.068) | -0.053 | <.001 |
| Psychological well-being: Via social support | 0.012 (0.002, 0.021) | 0.006 | .016 |
| Psychological well-being: Via loneliness then social support | -0.023 (-0.034, -0.012) | -0.013 | <.001 |
| Psychological well-being: Via personality | -0.777 (-0.979, -0.574) | -0.423 | <.001 |

***Note.*** *N = 2,417. B and its 95% confidence interval are in the fitted model-variable units. β is the fully standardized estimate. Intervals and two-sided p values use robust MLR inference; indirect and total associations use the delta method. Models adjusted for age using a four-degree-of-freedom natural spline, sex assigned at birth, race/ethnicity, education, marital status, insurance, everyday discrimination, medical discrimination, and food-insecurity risk.*

### Supplementary Table S8. Structural paths in personality models

| Association | B (95% CI) | β | p |
|---|---|---|---|
| **ACE-8 personality model** | | | |
| ACE-8 → Loneliness | 0.072 (0.035, 0.108) | 0.073 | <.001 |
| Loneliness → Social support | -0.514 (-0.552, -0.476) | -0.538 | <.001 |
| ACE-8 → Social support | -0.020 (-0.051, 0.011) | -0.021 | .212 |
| ACE-8 → General personality factor | -0.150 (-0.201, -0.099) | -0.127 | <.001 |
| Loneliness → Perceived stress | 0.307 (0.265, 0.349) | 0.315 | <.001 |
| Social support → Perceived stress | 0.007 (-0.031, 0.045) | 0.007 | .716 |
| General personality factor → Perceived stress | -0.329 (-0.364, -0.294) | -0.406 | <.001 |
| ACE-8 → Perceived stress | -0.008 (-0.037, 0.020) | -0.009 | .568 |
| Loneliness → Anxiety | 0.062 (0.014, 0.109) | 0.063 | .011 |
| Social support → Anxiety | 0.047 (0.002, 0.093) | 0.046 | .041 |
| General personality factor → Anxiety | -0.487 (-0.525, -0.450) | -0.601 | <.001 |
| ACE-8 → Anxiety | 0.011 (-0.022, 0.044) | 0.012 | .512 |
| Loneliness → Depression | 0.177 (0.130, 0.224) | 0.189 | <.001 |
| Social support → Depression | 0.039 (-0.002, 0.080) | 0.040 | .065 |
| General personality factor → Depression | -0.403 (-0.441, -0.366) | -0.517 | <.001 |
| ACE-8 → Depression | 0.039 (0.007, 0.070) | 0.042 | .016 |
| Loneliness → Psychological well-being | -0.505 (-0.608, -0.401) | -0.302 | <.001 |
| Social support → Psychological well-being | 0.224 (0.130, 0.319) | 0.128 | <.001 |
| General personality factor → Psychological well-being | 0.849 (0.733, 0.966) | 0.611 | <.001 |
| ACE-8 → Psychological well-being | 0.003 (-0.064, 0.070) | 0.002 | .919 |
| **ASRS-6 personality model** | | | |
| ASRS-6 → Loneliness | 0.178 (0.140, 0.217) | 0.179 | <.001 |
| Loneliness → Social support | -0.526 (-0.565, -0.487) | -0.550 | <.001 |
| ASRS-6 → Social support | 0.047 (0.015, 0.080) | 0.049 | .005 |
| ASRS-6 → General personality factor | -0.789 (-0.919, -0.659) | -0.536 | <.001 |
| Loneliness → Perceived stress | 0.304 (0.261, 0.347) | 0.309 | <.001 |
| Social support → Perceived stress | 0.006 (-0.032, 0.044) | 0.006 | .752 |
| General personality factor → Perceived stress | -0.284 (-0.325, -0.244) | -0.428 | <.001 |
| ASRS-6 → Perceived stress | 0.020 (-0.037, 0.076) | 0.020 | .496 |
| Loneliness → Anxiety | 0.065 (0.016, 0.114) | 0.066 | .010 |
| Social support → Anxiety | 0.047 (0.002, 0.092) | 0.046 | .042 |
| General personality factor → Anxiety | -0.421 (-0.463, -0.378) | -0.637 | <.001 |
| ASRS-6 → Anxiety | 0.006 (-0.059, 0.070) | 0.006 | .866 |
| Loneliness → Depression | 0.170 (0.124, 0.216) | 0.180 | <.001 |

| Association | B (95% CI) | β | p |
|---|---|---|---|
| Social support → Depression | 0.034 (-0.007, 0.075) | 0.034 | .102 |
| General personality factor → Depression | -0.338 (-0.377, -0.299) | -0.528 | <.001 |
| ASRS-6 → Depression | 0.077 (0.016, 0.139) | 0.082 | .014 |
| Loneliness → Psychological well-being | -0.547 (-0.663, -0.430) | -0.296 | <.001 |
| Social support → Psychological well-being | 0.245 (0.140, 0.350) | 0.127 | <.001 |
| General personality factor → Psychological well-being | 0.985 (0.818, 1.152) | 0.790 | <.001 |
| ASRS-6 → Psychological well-being | 0.388 (0.204, 0.572) | 0.211 | <.001 |

***Note.*** *N = 2,417. B and its 95% confidence interval are in the fitted model-variable units. β is the fully standardized estimate. Intervals and two-sided p values use robust MLR inference; indirect and total associations use the delta method. Models adjusted for age using a four-degree-of-freedom natural spline, sex assigned at birth, race/ethnicity, education, marital status, insurance, everyday discrimination, medical discrimination, and food-insecurity risk.*

### Supplementary Table S9. Incremental explained variance from social measures

| Exposure | Outcome | Both measures $R^2$ (%) | Add MOS-8 after ULS-8 $\Delta R^2$ (pp) | Add ULS-8 after MOS-8 $\Delta R^2$ (pp) |
|---|---|---|---|---|
| ACE-8 | Perceived stress | 51.58 | 0.14 | 10.73 |
| ACE-8 | Anxiety | 33.83 | 0.67 | 4.74 |
| ACE-8 | Depression | 39.30 | 0.48 | 7.61 |
| ASRS-6 | Perceived stress | 55.18 | 0.06 | 8.18 |
| ASRS-6 | Anxiety | 40.67 | 0.41 | 2.73 |
| ASRS-6 | Depression | 46.84 | 0.25 | 4.83 |

***Note.*** *N = 2,417. Each base model includes the exposure and the primary covariates. $\Delta R^2$ is the difference between nested-model $R^2$ values multiplied by 100, expressed in percentage points (pp).*

### Supplementary Table S10. Adjusted social measure coefficients across nested regressions

| Exposure and outcome | Model and predictor | B (95% CI) | β | p |
|---|---|---|---|---|
| ACE-8 / Perceived stress | Loneliness only / Loneliness | 0.446 (0.409, 0.482) | 0.444 | <.001 |
| ACE-8 / Perceived stress | Social support only / Social support | -0.194 (-0.235, -0.154) | -0.185 | <.001 |
| ACE-8 / Perceived stress | Both measures / Loneliness | 0.473 (0.432, 0.515) | 0.472 | <.001 |
| ACE-8 / Perceived stress | Both measures / Social support | 0.054 (0.013, 0.095) | 0.051 | .010 |
| ACE-8 / Anxiety | Loneliness only / Loneliness | 0.248 (0.203, 0.293) | 0.253 | <.001 |
| ACE-8 / Anxiety | Social support only / Social support | -0.045 (-0.091, 0.001) | -0.044 | .057 |
| ACE-8 / Anxiety | Both measures / Loneliness | 0.308 (0.258, 0.358) | 0.314 | <.001 |
| ACE-8 / Anxiety | Both measures / Social support | 0.117 (0.066, 0.167) | 0.114 | <.001 |
| ACE-8 / Depression | Loneliness only / Loneliness | 0.331 (0.288, 0.374) | 0.346 | <.001 |
| ACE-8 / Depression | Social support only / Social support | -0.103 (-0.148, -0.059) | -0.103 | <.001 |
| ACE-8 / Depression | Both measures / Loneliness | 0.381 (0.333, 0.429) | 0.397 | <.001 |
| ACE-8 / Depression | Both measures / Social support | 0.096 (0.050, 0.142) | 0.096 | <.001 |
| ASRS-6 / Perceived stress | Loneliness only / Loneliness | 0.402 (0.365, 0.438) | 0.401 | <.001 |
| ASRS-6 / Perceived stress | Social support only / Social support | -0.182 (-0.221, -0.143) | -0.174 | <.001 |
| ASRS-6 / Perceived stress | Both measures / Loneliness | 0.421 (0.380, 0.462) | 0.420 | <.001 |
| ASRS-6 / Perceived stress | Both measures / Social support | 0.036 (-0.003, 0.076) | 0.035 | .071 |
| ASRS-6 / Anxiety | Loneliness only / Loneliness | 0.190 (0.147, 0.233) | 0.193 | <.001 |
| ASRS-6 / Anxiety | Social support only / Social support | -0.032 (-0.075, 0.011) | -0.031 | .148 |
| ASRS-6 / Anxiety | Both measures / Loneliness | 0.238 (0.189, 0.286) | 0.242 | <.001 |
| ASRS-6 / Anxiety | Both measures / Social support | 0.092 (0.044, 0.139) | 0.089 | <.001 |
| ASRS-6 / Depression | Loneliness only / Loneliness | 0.272 (0.231, 0.314) | 0.284 | <.001 |
| ASRS-6 / Depression | Social support only / Social support | -0.091 (-0.132, -0.049) | -0.090 | <.001 |
| ASRS-6 / Depression | Both measures / Loneliness | 0.309 (0.263, 0.355) | 0.322 | <.001 |
| ASRS-6 / Depression | Both measures / Social support | 0.070 (0.026, 0.113) | 0.070 | .002 |

***Note.*** *N = 2,417. Models include the exposure and primary covariates. Confidence intervals and p values use HC3 standard errors with residual-degrees-of-freedom t inference. B is the model-variable units; β uses the predictor and outcome standard deviations in the analysis sample.*

### Supplementary Table S11. Loneliness and social support overlap diagnostics

| Exposure | Predictor | VIF | Tolerance | Pearson r | Partial r |
|---|---|---|---|---|---|
| ACE-8 | Loneliness | 2.076 | 0.482 | -0.647 | -0.519 |
| ACE-8 | Social support | 1.937 | 0.516 | -0.647 | -0.519 |
| ASRS-6 | Loneliness | 2.151 | 0.465 | -0.647 | -0.522 |
| ASRS-6 | Social support | 1.942 | 0.515 | -0.647 | -0.522 |

***Note.*** *N = 2,417. VIF includes the exposure and all primary covariates. Partial r residualizes both social measures on the same base predictors.*

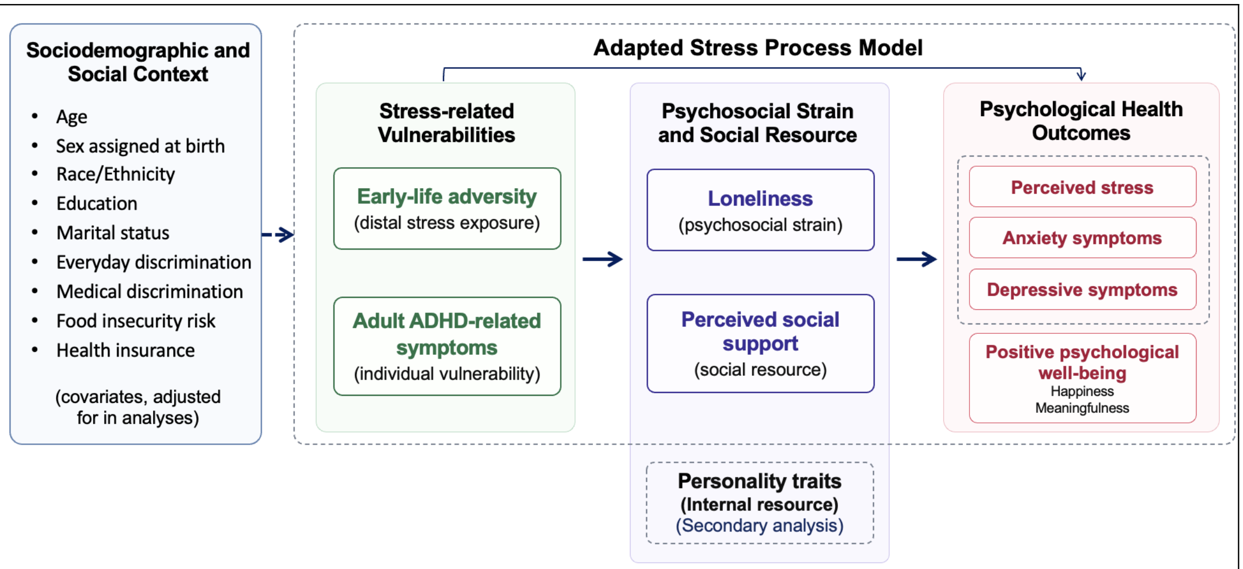


**Supplementary Figure S1. Adapted stress-process framework guiding the present study.** Guided by the Stress Process Model, the framework considers early-life adversity as a distal stress-related vulnerability and adult ADHD-related symptoms as an individual neurobehavioral vulnerability. Loneliness and perceived social support represent psychosocial strain and social resources, respectively, through which these vulnerabilities may be indirectly associated with perceived stress, anxiety symptoms, depressive symptoms, and psychological well-being. Direct associations between the primary stressors and psychological outcomes were also examined. Early-life adversity and adult ADHD-related symptoms were evaluated in separate parallel models. Personality characteristics were explored as additional individual characteristics in secondary analyses. Directional arrows represent theory-informed model specification and do not imply temporal or causal relationships.

**Abbreviations:** ACE-8, 8-item adverse childhood experiences score; ASRS-6, 6-item Adult ADHD Self-Report Scale; BFI-2-XS, 15-item Big Five Inventory-2 Extra-Short Form; GAD-7, 7-item Generalized Anxiety Disorder scale; IBS, irritable bowel syndrome; MOS-8, 8-item Medical Outcomes Study Social Support measure; PHQ-9, 9-item Patient Health Questionnaire; PSS-10, 10-item Perceived Stress Scale; ULS-8, 8-item UCLA Loneliness Scale.

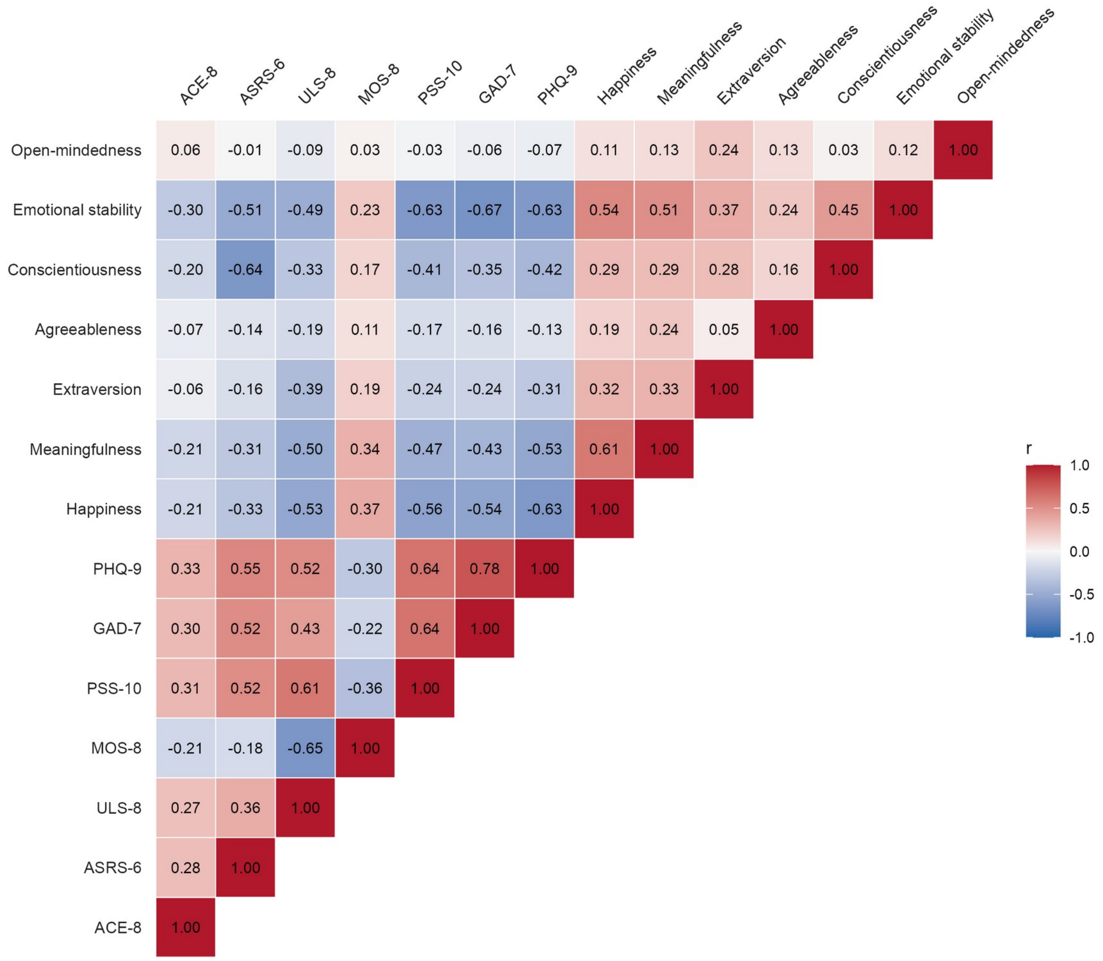


**Supplementary Figure S2. Pearson correlations among study variables.** Cell values are Pearson r for 2,417 participants. Red and blue indicate positive and negative correlations; shading represents their magnitude. ACE-8, adverse childhood experiences; ASRS-6, Adult ADHD Self-Report Scale; ULS-8, UCLA Loneliness Scale; MOS-8, Medical Outcomes Study social support; PSS-10, Perceived Stress Scale; GAD-7, Generalized Anxiety Disorder; PHQ-9, Patient Health Questionnaire. The five personality domains were measured using the BFI-2-XS.

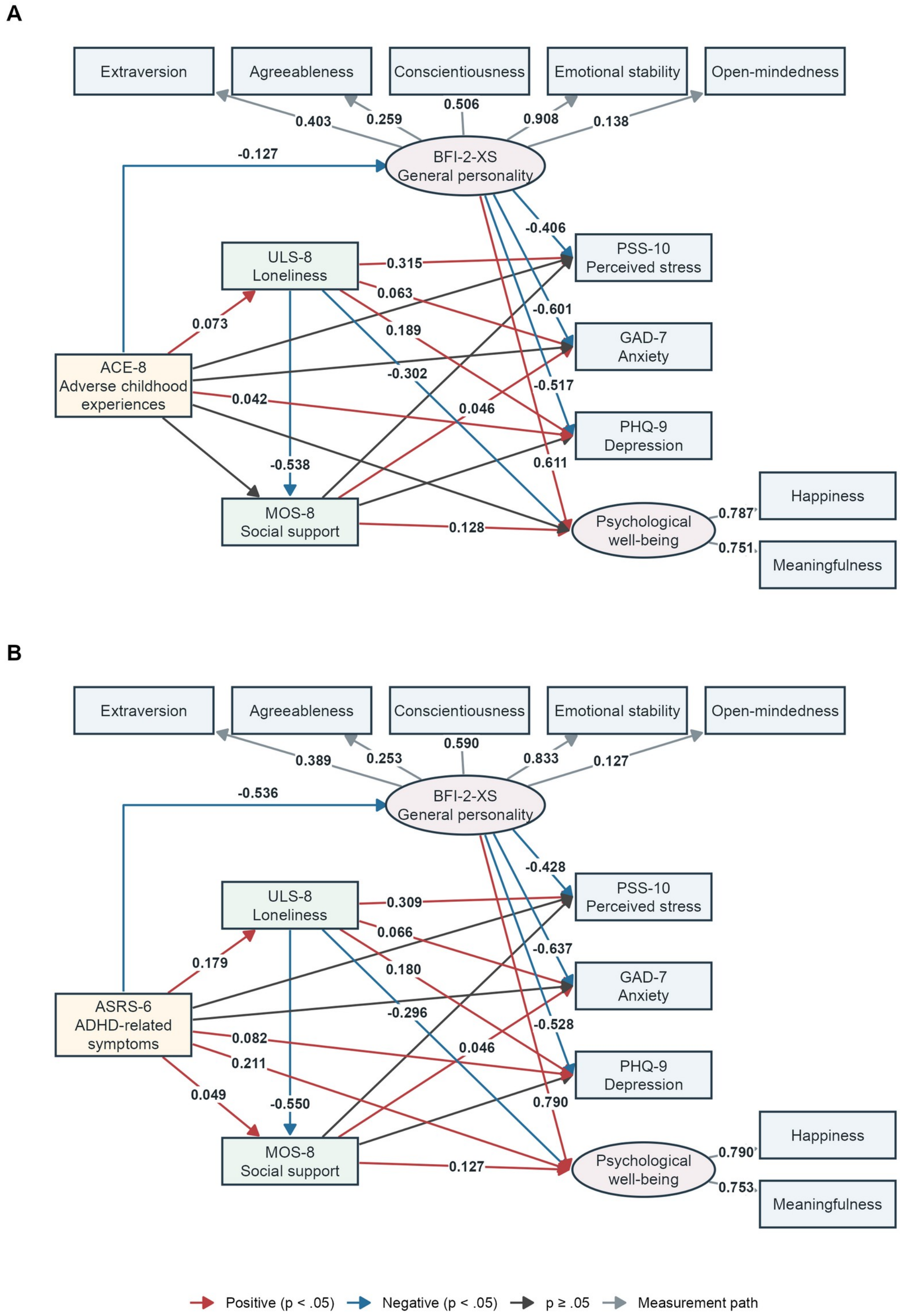


**Supplementary Figure S3. Exploratory personality-expanded structural equation models.** A, early-life adversity. B, ADHD-related symptoms. Ellipses indicate latent factors. Values are fully standardized coefficients for significant structural paths and measurement loadings. Red/blue arrows indicate positive/negative associations ($p < .05$), dark arrows indicate $p \geq .05$, and gray arrows indicate loadings. N = 2,417.
